# Spinal Anesthesia is Safe and Cost-effective for Laparoscopic Appendectomy in Children: A Case-Control Study

**DOI:** 10.1101/2020.10.20.20215798

**Authors:** Md Jafrul Hannan, Mosammat Kohinoor Parveenl, Alak Nandy, Md Samiul Hasan

## Abstract

**Background:** Owing to the widespread use of general anesthesia, administration of spinal anesthesia in pediatrics is not widely practiced. Yet there is ample positive evidence demonstrating its safety, effectiveness and success.

**Objective:** The objective of this study is to demonstrate that laparoscopic appendectomies are successful under spinal anesthesia and elicit clear advantages over general anesthesia.

**Methods:** This was a retrospective analysis of 77 pediatric (5-8 year old) laparoscopic appendectomies that took place in a Hospital in Chittagong, Bangladesh in 2019. Approximately half of the patients underwent spinal anesthesia while the other half underwent general anesthesia. Variables such as surgery and operation theatre times, pain score, incidence of post-surgery vomiting, analgesic usage, discharge times and hospital costs were recorded. Statistical analysis was used to analyze the data as a function of form of anesthesia.

**Results:** The probability of vomiting when using spinal compared to general anesthesia was much lower within the first 5 hours (P < .001) and after 6 hours (P = .008) of operation. Highly significant difference (P < .001) was observed in the total costs of the procedures. A significantly higher likelihood of patients being discharged the same day of the procedure was noted if spinal anesthesia was used (P = .008).

**Conclusions:** Spinal anesthesia is superior to general anesthesia for pediatric laparoscopic appendectomies. Patient comfort is improved through a significant decrease in vomiting. This enables more rapid hospital discharges and a significant cost saving, without compromising the outcome of the procedure.

**MINI-ABSTRACT:** Spinal anesthesia is seldom used for laparoscopy in children. This retrospective case-controlled study compared spinal anesthesia with general anesthesia in children between 5 and 8 years of age. Spinal anesthesia proved to be safer and cost-effective for laparoscopy in children.

## INTRODUCTION

The history and success of pediatric spinal anesthesia procedures, beginning with the 1898 report by Bier and adopted a few years later in several studies by Gray and Cantab has been recently documented.^1^ As general anesthesia technological developments improved, there was little interest in pediatric spinal anesthesia until the 1950s, when more studies advocated for use of spinal anesthesia in children.^2^ Since then, the spinal anesthetic approach has increased dramatically in children, and the potential problems and risks of general anesthesia in pediatrics documented.^3^ However, even by 1984 Abajian et al.^4^ noted that despite the reports of use of spinal anesthesia in children and confirmed it to be a safe alternative to general anesthesia, even for patients under one year of age, it remained underutilized. This group subsequently reported on their progress in 2006.^5^ Studies of their 1554 procedures found complication rates were very low, and they recommended spinal anesthesia for lower abdominal or extremity surgery in infants. An Italian and Finnish collaborative published^6^ studied 1132 children with a much wider age gap, between 6 months and 14 years, with similar conclusions (specifically with hyperbaric bupivacaine). In 2006, Imbelloni et al.^7^ reported on 307 consecutive cases of patients under the age of 13 years in a Brazilian setting and demonstrated an excellent rate of success, but cautioned that spinal anesthesia in children should be administered only by anesthesiologists already trained in spinal anesthesia in adults. They further noted that the cost to the facility was 54% less than the cost of general anesthesia, an important consideration in countries with limited financial resources. In Nigeria, even as recently as 2010, only general anesthesia was used. Their first study indicated that spinal anesthesia in children caused minimal hemodynamic disruption and was classified as a safe technique for lower extremity surgeries.^8^ In 2010, Polaner and Drescher^9^ and a year later, Ecoffey^10^ reviewed the safety record and came to the conclusion that although usage of regional anesthesia, whether as adjuncts, primary anesthesia or postoperative analgesia, was becoming increasingly common in pediatric practice, data on their safety remained limited because of the scarcity of large-scale prospective studies required to detect low incidence events. Despite this, their study concluded that regional blockades in infants and children appeared to have a very high degree of safety. They noted the importance of attention to technique, detail, and prudent patient selection to avoid possible complications.

Despite this positive evidence, even as recently as 2018, there have been some controversial topics regarding spinal anesthesia in pediatrics. The European Society of Regional Anesthesia and Pain Therapy/American Society of Regional Anesthesia and Pain Medicine published their recommendations on local anesthesia and adjuvant dosage in pediatric regional anesthesia, conspicuously noting that up to that point there was a large variability of dosages used in clinical practices. Their recommendations were intended to curb that variability.^11^ The technique is still gaining traction, and even as recently as 2019 its benefits have been again summarized^12^ while a recent report out of Pakistan^13^ noted that they have been performing surgeries successfully for the past 20 years using spinal anesthesia and the only real danger is when they were applied by poorly or untrained personnel.

Another recent area of debate is the applicability of spinal anesthesia to laparoscopic approaches to surgery. One of the first reports of laparoscopy under spinal anesthesia was reported by Islam et al.^14^ in 2014, where laparoscopic pyloromyotomy procedures in infants were investigated. Of the 12 cases studied, 9 were successful, while the other three cases required conversion to general anesthesia. The three failures were related to the inability to access the intrathecal space and an inadequate block level so that the infant did not tolerate insufflations of the abdomen. More recently, Chiao and Boretsky^15^ presented three case reports employing laparoscopic surgery for inguinal hernia repair. All procedures were successful, with one patient experiencing hypertension and tachycardia during insufflations with brief supplemental use of sevoflurane. The authors concluded that the use of spinal anesthesia for laparoscopic surgery was successful, with the advantage of decreased exposure to opioids and general anesthesia agents, some of which are potential neurotoxins that may negatively affect brain development. This can provide an additional anesthesia option for providers and families. They finally claim that laparoscopy could, perhaps, no longer be viewed as being incompatible with the use of spinal anesthesia in infants.

Despite the increased prevalence and positive outlooks of spinal anesthesia in children, it is still not practiced everywhere owing to the widespread use of conventional general anesthesia. In this article, we will demonstrate through a retrospective study of 77 patients, that laparoscopic appendectomies can be performed quite successfully under spinal anesthesia and investigate the effects, if any, on patient comfort measured by the incidence of vomiting in the postoperative period. Furthermore, we will provide clear evidence of other clinical advantages of using the spinal technique compared to general anesthesia, including significantly lower hospital costs and quicker hospital discharges.

## METHODS

This study was a retrospective analysis of 77 pediatric (5-8 year old children) laparoscopic appendectomies that took place in the South Point Hospital, Chittagong, Bangladesh between January 1, 2019 and December 31, 2019. The ethical clearance for this study was provided by the South Point Hospital (Admn/SPH/191/2020). There was a roughly even distribution between patients undergoing spinal and general anesthesia (Table 1). Those receiving spinal anesthesia also received sedation with diazepam or ketamine hydrochloride injection as an adjunct to alleviate their anxiety and help them remain calm. Patients who received general anesthesia also received nitrous oxide gas throughout the intraoperative period as analgesics and were kept relaxed by norcuronium.

| Demographic Data |  |  |  |
| --- | --- | --- | --- |
| Gender |  | n | % |
| Female |  | 38 | 49.4 |
| Male |  | 39 | 50.6 |
| Age |  |  |  |
| Age 5 |  | 17 | 17.2 |
| Age 6 |  | 17 | 17.2 |
| Age 7 |  | 24 | 24.3 |
| Age 8 |  | 19 | 19.3 |
| Anesthesia Type |  |  |  |
| Spinal Anesthesia |  | 40 | 52.0 |
| General Anesthesia |  | 37 | 48.0 |

Spinal anesthesia consisted of 0.5% bupivacaine in 8.5% dextrose at a dose of 0.4 mg/kg of body weight. CO2 insufflations pressures were kept under 8 mmHg and the flow was maintained between 2.0-2.5 litres/minute. For all procedures, irrespective of the type of anesthesia, antiemetics were administered at the start of the procedure, while dosages of NSAIDs were administered towards the end of the operation. This is the usual practice in hospitals. Feeding was recommenced 4-5 hours postoperatively for the general anesthesia group and 2-3 hours postoperatively in the spinal anesthesia group. Visual Analog Scale (VAS) pain scores were recorded at 1, 3, and 6 hours postoperatively in all cases.

Statistical analysis of the data was performed using JMP statistical software (SAS, Cary NC, USA). Significance was held at the 95% level unless otherwise noted (minimum 90% level). Chi-squared and Fisher’s Exact Test were used for contingency analysis of categorical data.

Parametric (Student -t tests) or non-parametric tests (Wilcoxon) were used for comparison of continuous numerical data depending on the normality of the data, determined using the Shapiro-Wilk test. The effects of anesthesia on vomiting during the first five hours post-operation, after six hours post-operation, the speed of patient discharge, and the cost of the procedure were examined. Attempts at associating pain scores and analgesic requirements after the procedure to the two forms of anesthesia were also made. However, the administration of NSAIDs intraoperatively, and the use of different adjuncts during the procedures present confounding factors that could mask the true pain score and analgesic requirements.

## RESULTS

The descriptive statistics for the cohort of 77 patients are presented in Table 1. The data indicate an approximate even distribution of patients across gender, age, and anesthesia method used for the procedure.

The results of the incidence of vomiting analysis are shown by the mosaic plots in Figure 1, which may be thought of as a visual representation of a contingency table. In these plots, the column width is proportional to n, and the height of each section represents the fraction of data within that category. The narrow band on the right side represents the distribution across the entire group. For both time points, the probability of vomiting was significantly lower for spinal anesthesia, a result confirmed by Fisher’s Exact Test (P< .001 for vomiting within the first 5 hours and P = .008 for vomiting after 6 hours).

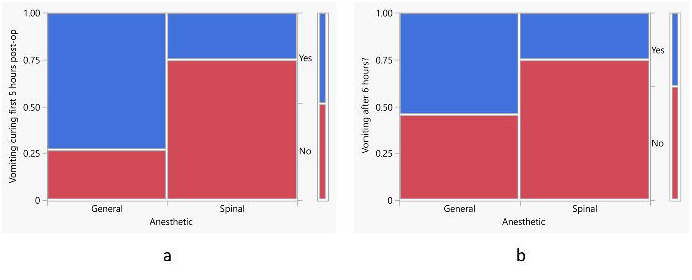

Some gender-related effects also presented themselves. Figure 2 shows the mosaic plots for the incidence of vomiting within the first five hours of the procedure. In both cases, there was a significantly greater likelihood of vomiting when general anesthesia was used, but it was far more significant for the male population (Fisher’s Exact Test; P< .001) than for females (P = .04). Six hours after the operation, the females still displayed a significantly greater likelihood of vomiting if general anesthesia was used during the procedure (P = .02). However, this effect disappeared for the male group, and the incidence of vomiting was independent of the anesthesia type (P = .32).

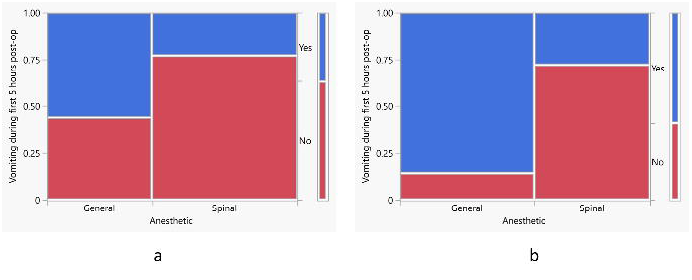

We also observed some interesting effects when we binned the data into a “younger” group consisting of the 5-6 year olds and an “older” group consisting of the 7-8 year olds. In terms of vomiting within the first 5 hours of the procedure, both groups showed a significant increase in the likelihood of vomiting if general anesthesia was used (P = .02) for the younger group; P < .001 for the older group). After 6 hours, the incidence of vomiting remained significantly higher in the younger group (P = .04); however, for the older group, this incidence was independent of the type of anesthesia used (P =.12).

The antiemetic and NSAID administered during all procedures, irrespective of anesthesia type, followed the same regimen and is standard practice for this clinic. However, different adjuncts and sedatives administered intraoperatively will introduce confounding factors, making interpretation difficult. The results are presented in the Supplemental Content. The pain scores for 1 and 6 hours after the procedure were determined to be independent of the anesthesia type (two-sided P = .36 and .65, respectively – See Figure, Supplemental Content 1, showing mosaic plots for the pain scores comparing the two forms of anesthesia). At 3 hours post-surgery, a faint correlation observed at the 90% level of significance weakly suggests that higher pain scores were more likely to be observed when general anesthesia was used (one-sided P = .07). When stratified by gender or age bracket, this observation was highly significant for males (P = .02) and the older age bracket (P = .03), respectively (see Table, Supplemental Content 2, which summarizes the main findings of effect of anesthesia type on pain scores 1-, 3- and 6-hours post operation.). There was no association between age group and gender (P = .82; two-sided), thereby excluding a male-dominated result. Similar results, not too surprisingly, were found for analgesic administration. The analgesic administrations at 1 and 6-hours post operation were found to be the same regardless of the type of anesthesia. At 3-hours post operation a weak association between anesthesia and analgesic use was observed at the 90% level of significance (P = .06) for males, while a more significant association was observed for the7-8 year old cohort of patients at the 95% level of significance (P = .04). The mosaic plots (see Figure, Supplemental Content 3, which presents analgesic usage as a function of anesthesia at 3-hours post operation for males and the 7-8 year old age bracket.) suggest that the association is related to a substantial tendency for those utilizing spinal anesthesia that does not require an analgesic.

The second goal of the study was to investigate the cost reduction for hospitals that might be achieved using spinal anesthesia procedures. These data are presented in Figure 3 (note one data point for the cost was missing in the spinal anesthesia category). A highly significant difference, confirmed by a Wilcoxan test (P < .001), was observed, with the total cost of spinal anesthesia procedures being approximately one-fifth the cost of a typical general anesthesia procedure.

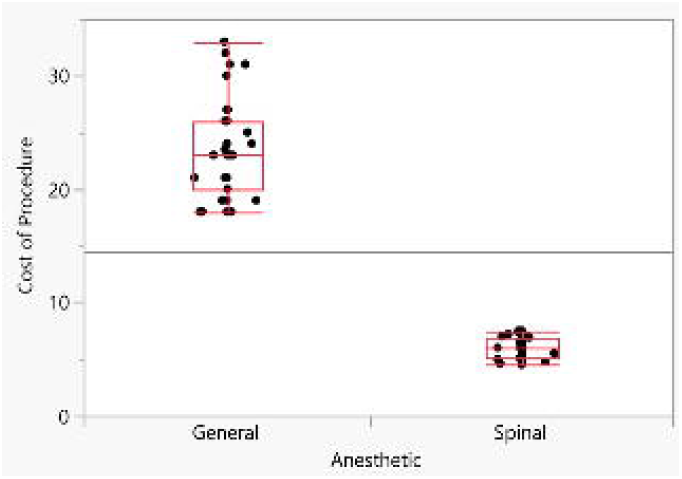

We were also interested in whether any difference would be found in the expediency of discharge as a function of anesthesia form. The results were also encouraging, as illustrated by the mosaic plot in Figure 4, which indicates a significantly higher likelihood of patients being discharged the same day of the procedure if a spinal anesthesia was used (Fisher’s Exact Test; P = .008).

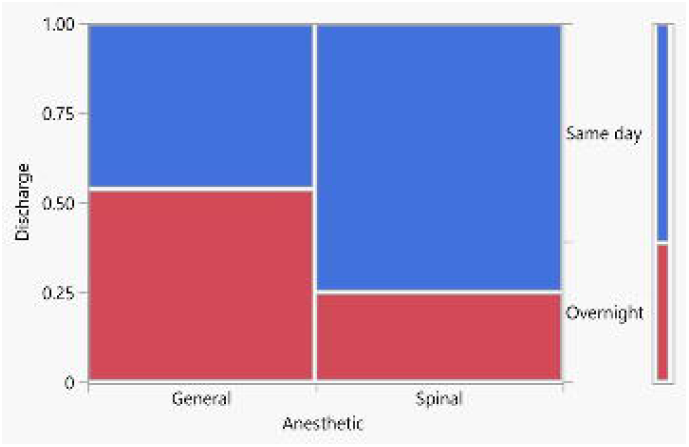

Gender was also significant in discharge times. The probability that female patients would be discharged on the same day was significantly higher if spinal anesthesia was used (Fisher’s Exact Test; P = .04), while for the male population there was no real effect of anesthesia type in terms of discharge time (P =.11).

There was also an age effect observed in the discharge time. For the younger group, those who received spinal anesthesia were far more likely to be discharged on the same day of the procedure (P = .02), while for the older group, this was not the case and discharge time was independent of anesthesia mode (P = .18). A summary of the main findings from this study is presented in Table 2.

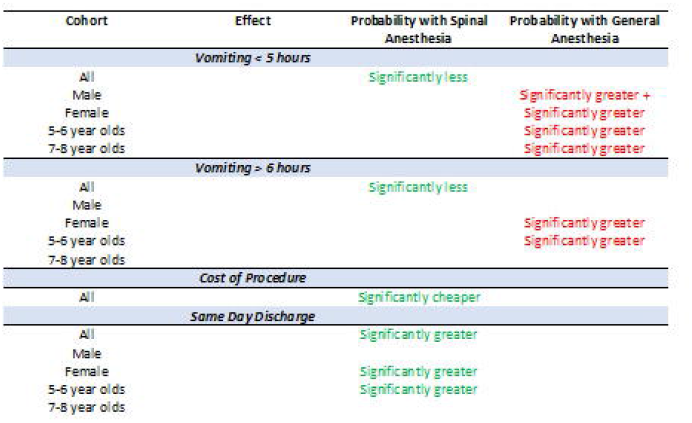

The effects of the adjuncts diazepam and ketamine hydrochloride on the spinal anesthesia group were also examined in terms of incidence of vomiting, but no significant differences were found up to 5 hours postoperatively (Fisher’s Exact Test, two-tailed; P = .26) or after 6 hours postoperatively (P = .48). These adjuncts also did not affect the cost of the procedure (Student’s t-test; P = .26) nor the speed of discharge (Fisher’s Exact Test, two-tailed; P = .48).

Finally, all factors were combined in a multiple correspondence analysis (MCA). Multiple correspondence analyses are the categorical equivalent of principal component analysis in multivariate statistics. It produces a plot which is a 2-dimensional representation of “n-space” where n is the number of variables. The two dimensions chosen are those that explain the most variance in the data. The closer the points are to this plot, the more highly they are associated with one another on a relative basis, while the further away from the origin the points are located, the more they are discriminating themselves from the rest of the data. Here, the operation time and the theatre time were binned into two categories: above and below the median value. The cost of the procedure has been binned into “less expensive” (less than 15k) and “more expensive” (greater than 15k) categories. The resultant plot is shown in Figure 5, where the top portion shows the entire MCA plot, while the bottom zooms in the area where the “less expensive” category is located. The plot of these two dimensions explains 32% of the variance in the data and shows astonishingly well how “less expensive” and spinal anesthesia are associated. Other factors found to be associated with the less expensive category included an operation theatre time between and 25-40 minutes (the shortest time bin), no vomiting during the first 5 hours or after 6 hours post operation, and same-day discharge. These factors all make sense, but other interesting factors include the use of NSAIDs as analgesics 1-hour post operation as well as use of either of the two adjuncts, but that is expected as they were only used with spinal anesthesia.

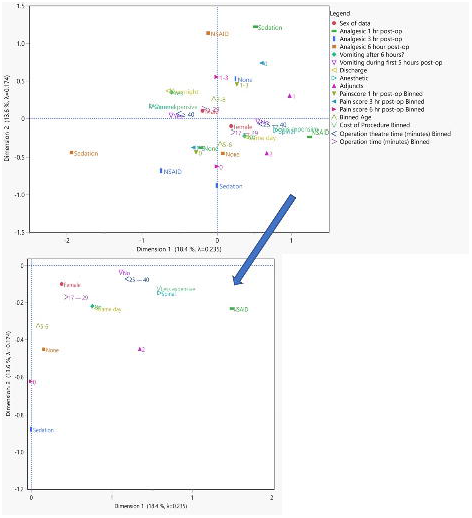

## DISCUSSION

Laparoscopic surgery is now the method of choice for lower abdominal and extremity procedures. Childers et al.^16^ reported that 9507 appendectomies in children under the age of 18 years in the USA, 94.6% were conducted using laparoscopy. In four central European institutions, of the 519 pediatric appendectomies performed, 79.6% were conducted via laparoscopy.^17^ In Germany; Gosemann et al.^18^ found that of 8110 pediatric appendectomies, 75% were performed using laparoscopy. In 2018, in a wide-ranging study, Tom et al.^19^ found that of the 58511 appendectomies conducted in children’s hospitals in the USA between 2003 and 2012, 70% were done using laparoscopy, compared to 53% of the ∼1.2 million conducted at non-children’s hospitals. Zani et al.^20^ summarized the results of the European Pediatric Surgeons’ Association survey on the management of pediatric appendicitis, compiled from 169 respondents from 42 countries (24 European). For simple appendicitis, laparoscopy was the preferred method for 89%, while for perforated appendicitis, it remained the method of choice for 81%. In Japan, Fujishiro et al.^21^ found that of the 4489 pediatric appendectomies performed, 70.5% were performed laparoscopically. It is clear from these studies that for pediatric appendicitis, laparoscopy is the method of choice, and also concluded in a review of pediatric appendicitis by Rentea et al.^22^ However, in all of these significant studies, an important fact is conspicuously absent. No mention of the type of anesthesia administered during the procedure is given. An Egyptian study of 390 pediatric complicated appendicitis cases was published by Khirallah et al.^23^ comparing laparoscopic (200 cases) versus open appendectomies. All procedures were conducted under general anesthesia and concluded that the laparoscopic technique should be the first choice of procedure for pediatric surgeons for appendectomy procedures. Thus, the present study clearly addresses a paucity of data pertaining to the effect of the type of anesthesia on pediatric laparoscopic procedures in terms of post-operative patient vomiting, discharge rate, and relative costs.

The results of the present study clearly showed that the use of spinal anesthesia reduced the likelihood of vomiting during both the first five hours and six hours postoperatively (Figures 1 and 2 and Table 2). This mirrors the results of Verma et al.^24^ in their study of 102 pediatric patients aged 6 months to 14 years undergoing various surgeries including herniotomy, appendectomy, genitourinary surgeries, and lower limb orthopedic surgeries under spinal anesthesia. In this cohort, no incidence of vomiting was noted. Similarly, Ahmed et al.^25^ in their study of 78 children with a similar range of procedures reported 6 cases of nausea and 1 case of vomiting. Kokki and Hendolin.^26^ in a cohort of 52 patients between the ages of 7 and 18 years undergoing lower umbilical procedures with spinal anesthesia (Bupivacaine in 8% dextrose) reported 10 patients experiencing nausea but no vomiting. None of these studies stratified the incidence of vomiting by gender, so in that respect the results of this study are, to the best of our knowledge, novel. However, Verma et al.^24^ and Ahmed et al.^25^ studies were largely male-dominated (> 80%); therefore, the observation in Figure 2 that males are especially less likely to experience vomiting in the first 5 hours post-operatively is not unexpected. There is also a weak suggestion that there may be a relationship between pain scores and anesthesia at the 3-hour postoperative period, which becomes more significant for males and the 7-8 year old age bracket. However, confounding factors from the different adjuncts delivered intraoperatively make these results somewhat more difficult to interpret. In fact, the entire subject of post-operative nausea and vomiting can be quite complex.^27^

There is ample evidence for shorter hospital stays with a laparoscopic procedure,^18,28–30^ although the Japanese study^21^ contradicted this observation. They found no significant difference between laparoscopic and open appendectomies in terms of length of stay. The present results show a definite trend to overnight stays when general anesthesia is used, whereas same-day discharges are more highly associated with spinal anesthesia (Figure 4). This is especially true for females.

Teja et al.^31^ have championed the need for more cost-effectiveness research in anesthesiology. They noted a paucity of cost-effectiveness data, and even more so from a pediatric perspective. Although the research to this end is relatively simplistic and relates only to the cost of the procedures, a significant reduction in cost was found (by almost a factor of 5, Figure 3) in this study when spinal anesthesia was used. Imbelloni et al.^7^ reported a savings in anesthesia cost of 54% when the spinal method was used compared to their historical data. This was pooled over a wide variety of pediatric procedures.

In summary, the results of this study provide a clear indication that spinal anesthesia is superior to general anesthesia. The data provide strong evidence for more rapid hospital discharges and a significant cost saving, without compromising the outcome of the procedure. Patient comfort is also improved through a significant decrease in the lack of vomiting and possible lower pain scores.

## Supporting information

C:\Users\Dell\Documents\add pub\Prep manuscript\lap spinal-kolabtree appendectomy\medRxiv revision\Supplemental Digital Content 1.doc

C:\Users\Dell\Documents\add pub\Prep manuscript\lap spinal-kolabtree appendectomy\medRxiv revision\Supplemental Digital Content 2.doc

C:\Users\Dell\Documents\add pub\Prep manuscript\lap spinal-kolabtree appendectomy\medRxiv revision\Supplemental Digital Content 3.doc

## Data Availability

The datasets generated during and/or analyzed during the current study are available at Figshare

https://doi.org/10.6084/m9.figshare.13093541.v1

## ACKNOWLEDGMENTS

None

## AUTHOR CONTRIBUTIONS

All four authors have designed the study, performed the experiments, analyzed the data and wrote the manuscript.

## CONFLICT OF INTEREST STATEMENT

The authors have declared that no competing interests exist. ETHICS STATEMENT

The study was approved by the Ethics Committee of South Point Hospital.

## FUNDING STATEMENT

The funders had no role in study design, data collection and analysis, decision to publish, or preparation of the manuscript.

## DATA AVAILABILITY STATEMENT

The datasets generated during and/or analyzed during the current study are available at https://doi.org/10.6084/m9.figshare.13093541.v1.

## Notes

### Competing Interest Statement

The authors have declared no competing interest.

### Author Declarations

The ethical clearance for this study was provided by the South Point Hospital (Admn/SPH/191/2020).

### Summary of Updates

Changed some author information.

## REFERENCES

1. Brown TCK. History of pediatric regional anesthesia. Pediatr Anesth. 2012;22(1):3–9. doi:10.1111/j.1460-9592.2011.03636.x

2. Berkowitz S, Greene BA. Spinalanesthesia in children. Anesthesiology. 1951;12(3):376–387. doi:10.1097/00000542-195105000-00013

3. Becke K. Complications in pediatric anesthesia (Komplikationen in der Kinderanästhesie). Anaesthesist. 2014;63(7):548–554. doi:10.1007/s00101-014-2357-0

4. Abajian JC, Mellish RWP, Browne AF, Perkins FM, Lambert DH, Mazuzan Jr JE. Spinal Anesthesia for Surgery in the High-Risk Infant. Anesth Analg. 1984;63:359–362.

5. Williams RK, Adams DC, Aladjem E V., et al. The Safety and Efficacy of Spinal Anesthesia for Surgery in Infants: The Vermont Infant Spinal Registry. Anesth Analg. 2006;102(1):67–71. doi:10.1213/01.ANE.0000159162.86033.21

6. Puncuh F, Lampugnani E, Kokki H. Use of spinal anaesthesia in paediatric patients: a single centre experience with 1132 cases. Pediatr Anesth. 2004;14(7):564–567. doi:10.1111/j.1460-9592.2004.01240.x

7. Imbelloni LE, Vieira EM, Sperni F, Guizellini RH, Tolentino AP. Spinal anesthesia in children with isobaric local anesthetics: Report on 307 patients under 13 years of age. Pediatr Anesth. 2006;16(1):43–48. doi:10.1111/j.1460-9592.2005.01680.x

8. Rukewe A, Alonge T, Fatiregun A. Spinal anesthesia in children: no longer an anathema! Pediatr Anesth. 2010;20(11):1036–1039. doi:10.1111/j.1460-9592.2010.03431.x

9. Polaner DM, Drescher J. Pediatric regional anesthesia: what is the current safety record? Pediatr Anesth. 2011;21(7):737–742. doi:10.1111/j.1460-9592.2010.03499.x

10. Ecoffey C. Safety in pediatric regional anesthesia. Pediatr Anesth. 2012;22(1):25–30. doi:10.1111/j.1460-9592.2011.03705.x

11. Suresh S, Ecoffey C, Bosenberg A, et al. The European Society of Regional Anaesthesia and Pain Therapy/American Society of Regional Anesthesia and Pain Medicine Recommendations on Local Anesthetics and Adjuvants Dosage in Pediatric Regional Anesthesia. Reg Anesth Pain Med. Published online January 2018:1. doi:10.1097/AAP.0000000000000702

12. Whitaker EE, Williams RK. Epidural and Spinal Anesthesia for Newborn Surgery. Clin Perinatol. 2019;46(4):731–743. doi:10.1016/j.clp.2019.08.007

13. Durrani H-D. Pediatric spinal anesthesia at D.G. Khan (Pakistan); Our experience of 20 years. Anaesthesia, Pain Intensive Care. 2020;24(1):115. doi:10.35975/apic.v24i1.1238

14. Islam S, Larson SD, Kays DW, Irwin MD, Carvallho N. Feasibility of laparoscopic pyloromyotomy under spinal anesthesia. J Pediatr Surg. 2014;49(10):1485–1487. doi:10.1016/j.jpedsurg.2014.02.083

15. Chiao F, Boretsky K. Laparoscopic Surgery in Infants Under Spinal Anesthesia Block. A A Pract. 2019;12(5):168–170. doi:10.1213/XAA.0000000000000876

16. Childers CP, Dworsky JQ, Massoumi RL, et al. The contemporary appendectomy for acute uncomplicated appendicitis in children. Surgery. 2019;165(5):1027–1034. doi:10.1016/j.surg.2018.12.019

17. Dotlacil V, Frybova B, Polívka N, et al. Current management of pediatric appendicitis: A Central European survey. Adv Clin Exp Med. 2020;29(6):745–750. doi:10.17219/acem/122176

18. Gosemann J-H, Lange A, Zeidler J, et al. Appendectomy in the pediatric population—a German nationwide cohort analysis. Langenbeck’s Arch Surg. 2016;401(5):651–659. doi:10.1007/s00423-016-1430-3

19. Tom CM, Won RP, Lee AD, Friedlander S, Sakai-Bizmark R, Lee SL. Outcomes and Costs of Common Surgical Procedures at Children’s and Nonchildren’s Hospitals. J Surg Res. 2018;232:63–71. doi:10.1016/j.jss.2018.06.021

20. Zani A, Hall N, Rahman A, et al. European Paediatric Surgeons’ Association Survey on the Management of Pediatric Appendicitis. Eur J Pediatr Surg. 2019;29(01):053–061. doi:10.1055/s-0038-1668139

21. Fujishiro J, Watanabe E, Hirahara N, et al. Laparoscopic Versus Open Appendectomy for Acute Appendicitis in Children: a Nationwide Retrospective Study on Postoperative Outcomes. J Gastrointest Surg. Published online March 3, 2020. doi:10.1007/s11605-020-04544-3

22. Rentea RM, Peter SD St. Snyder CL. Pediatric appendicitis: state of the art review. Pediatr Surg Int. 2017;33(3):269–283. doi:10.1007/s00383-016-3990-2

23. Khirallah MG, Eldesouki NI, Elzanaty AA, Ismail KA, Arafa MA. Laparoscopic versus open appendectomy in children with complicated appendicitis. Ann Pediatr Surg. 2017;13(1):17–20. doi:10.1097/01.XPS.0000496987.42542.dd

24. Verma D, Naithani U, Gokula C, Harsha. Spinal anesthesia in infants and children: A one year prospective audit. Anesth Essays Res. 2014;8(3):324. doi:10.4103/0259-1162.143124

25. Ahmed M, Ali N, Kabir S, Nessa M. Spinal Anaesthesia: Is it Safe in Younger Children? J Armed Forces Med Coll Bangladesh. 1970;6(1):25–28. doi:10.3329/jafmc.v6i1.5988

26. Kokki H, Hendolin H. Hyperbaric bupivacaine for spinal anaesthesia in 7-18 yr old children: comparison of bupivacaine 5 mg ml-1 in 0.9% and 8% glucose solutions. Br J Anaesth. 2000;84(1):59–62. doi:10.1093/oxfordjournals.bja.a013382

27. Borgeat A, Ekatodramis G, Schenker CA. Postoperative Nausea and Vomiting in Regional Anesthesia. Anesthesiology. 2003;98(2):530–547. doi:10.1097/00000542-200302000-00036

28. Grewal H, Sweat J, Vazquez WD. Laparoscopic Appendectomy in Children Can Be Done as a Fast-Track or Same-Day Surgery. JSLS. 2004;8:151–154.

29. Jen HC, Shew SB. Laparoscopic Versus Open Appendectomy in Children: Outcomes Comparison Based on a Statewide Analysis. J Surg Res. 2010;161(1):13–17. doi:10.1016/j.jss.2009.06.033

30. Lee SL. Laparoscopic vs Open Appendectomy in Children. Arch Surg. 2011;146(10):1118. doi:10.1001/archsurg.2011.144

31. Teja BJ, Sutherland TN, Barnett SR, Talmor DS. Cost-Effectiveness Research in Anesthesiology. Anesth Analg. 2018;127(5):1196–1201. DOI: 10.1213/ANE.0000000000003334

