## Supplementary material for "Spinal Anesthesia is Safe and Cost-effective for Laparoscopic Appendectomy in Children: A Case-Control Study": C:\Users\Dell\Documents\add pub\Prep manuscript\lap spinal-kolabtree appendectomy\medRxiv revision\Supplemental Digital Content 1.doc

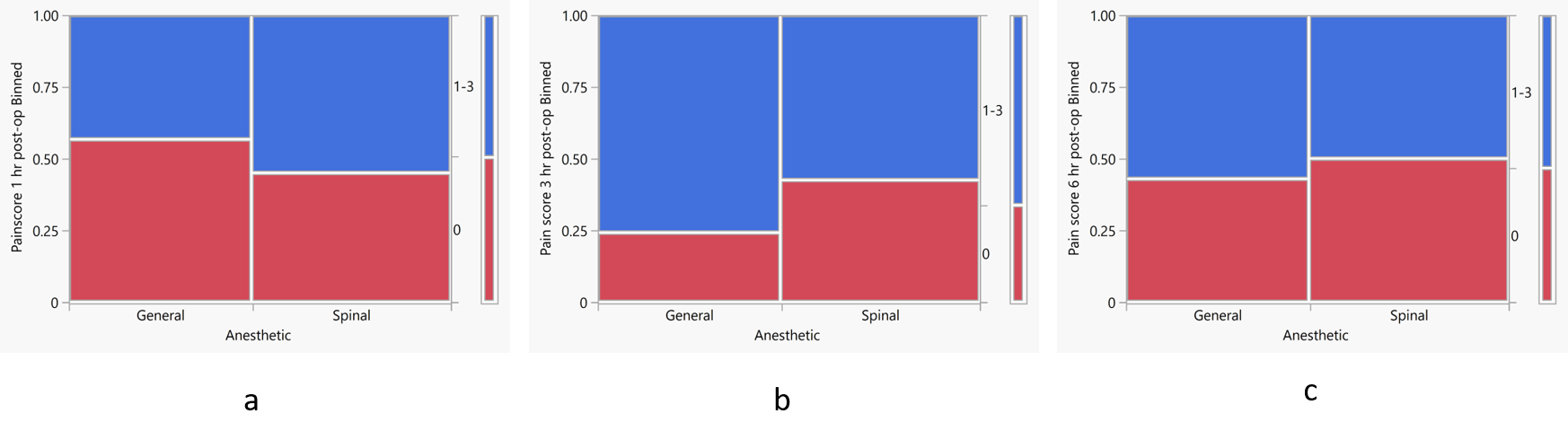

Supplemental Digital Content 1: Pain scores recorded (a) 1 hour, (b) 3 hours and (c) 6 hours post-operatively as a function of anesthesia. The score values are binned into two levels: 0 (no pain) and 1-3 (mild to severe pain).
