## Supplementary material for "Spinal Anesthesia is Safe and Cost-effective for Laparoscopic Appendectomy in Children: A Case-Control Study": C:\Users\Dell\Documents\add pub\Prep manuscript\lap spinal-kolabtree appendectomy\medRxiv revision\Supplemental Digital Content 2.doc

| ***Cohort*** | ***1 hour Pain Score*** | ***Probability with Spinal Anesthesia*** | ***Probability with General Anesthesia*** |
| --- | --- | --- | --- |
| All cohorts | No effect of anesthetic | | |
|  | ***3 hour Pain Score*** |  |  |
| All |  |  | Weakly higher |
| Male |  |  | Significantly higher |
| Female |  |  |  |
| 5-6 year olds |  |  |  |
| 7-8 year olds |  |  | Significantly higher |
|  | ***6 hour Pain Score*** |  |  |
| All cohorts | No effect of anesthetic | | |

Supplemental Digital Content 2: Summary of main findings of effect of anesthesia type on pain scores 1-, 3- and 6-hours post operation.
