## Supplementary material for "Spinal Anesthesia is Safe and Cost-effective for Laparoscopic Appendectomy in Children: A Case-Control Study": C:\Users\Dell\Documents\add pub\Prep manuscript\lap spinal-kolabtree appendectomy\medRxiv revision\Supplemental Digital Content 3.doc

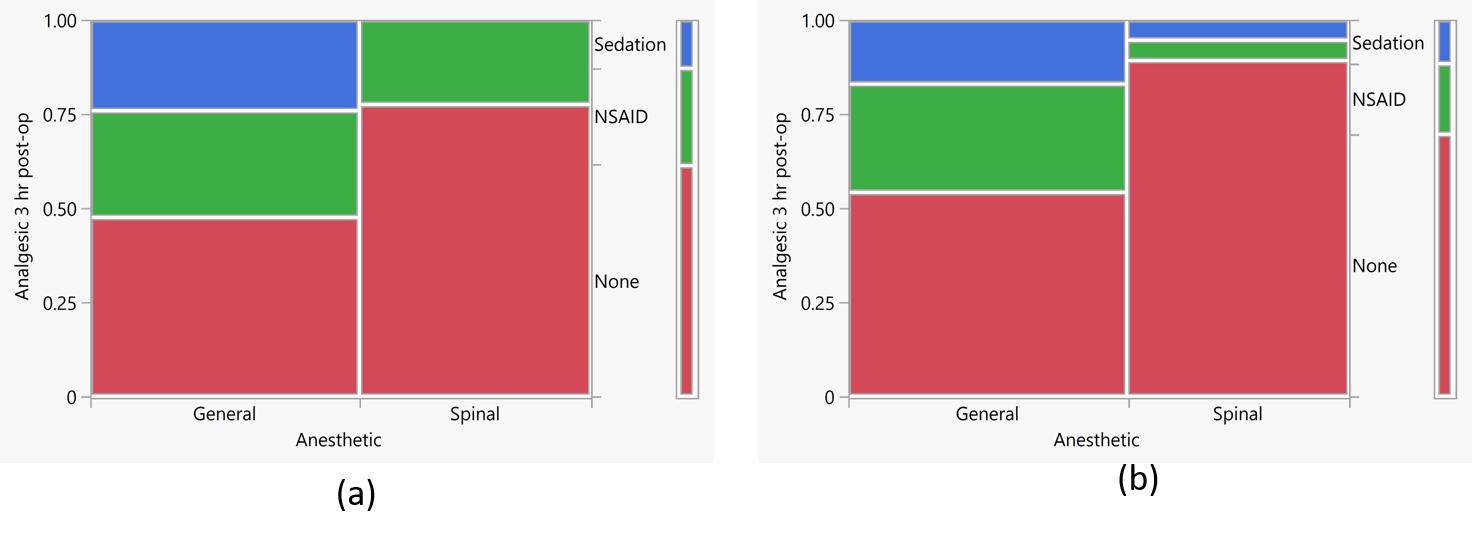

Supplemental Digital Content 3: Mosaic plots showing analgesic usage as a function of anesthesia at 3-hours post operation for (a) males and (b) the 7-8 year old age bracket.
